# A genome-wide association study suggests new susceptibility loci for primary antiphospholipid syndrome

**DOI:** 10.1101/2023.12.05.23299396

**Authors:** Desiré Casares-Marfil, Manuel Martínez-Bueno, Maria Orietta Borghi, Guillermo Pons-Estel, PRECISESADS Clinical Consortium, Guillermo Reales, Yu Zuo, Gerard Espinosa, Timothy Radstake, Lucas L. van den Hoogen, Chris Wallace, Joel Guthridge, Judith A James, Ricard Cervera, Pier Luigi Meroni, Javier Martin, Jason S. Knight, Marta E. Alarcón-Riquelme, Amr H. Sawalha

## Abstract

**Objectives:** Primary antiphospholipid syndrome (PAPS) is a rare autoimmune disease characterized by the presence of antiphospholipid antibodies and the occurrence of thrombotic events and pregnancy complications. Our study aimed to identify novel genetic susceptibility loci associated with PAPS.

**Methods:** We performed a genome-wide association study comprising 5,485 individuals (482 affected individuals) of European ancestry. Significant and suggestive independent variants from a meta-analysis of approximately 7 million variants were evaluated for functional and biological process enrichment. The genetic risk variability for PAPS in different populations was also assessed. Hierarchical clustering, Mahalanobis distance, and Dirichlet Process Mixtures with uncertainty clustering methods were used to assess genetic similarities between PAPS and other immune-mediated diseases.

**Results:** We revealed genetic associations with PAPS in a regulatory locus within the HLA class II region near *HLA-DRA* and in *STAT4* with a genome-wide level of significance. 34 additional suggestive genetic susceptibility loci for PAPS were also identified. The disease risk allele in the HLA class II locus is associated with overexpression of *HLA-DRB6*, *HLA-DRB9*, *HLA-DPB2*, *HLA-DQA2* and *HLA-DQB2*, and is independent of the association between PAPS and *HLA-DRB1*1302*. Functional analyses highlighted immune and nervous system related pathways in PAPS-associated loci. The comparison with other immune-mediated diseases revealed a close genetic relatedness to neuromyelitis optica, systemic sclerosis, and Sjögren’s syndrome, suggesting colocalized causal variations close to *STAT4*, *TNPO3*, and *BLK*.

**Conclusions:** This study represents a comprehensive large-scale genetic analysis for PAPS and provides new insights into the genetic basis and pathophysiology of this rare disease.

## 1. INTRODUCTION

Antiphospholipid syndrome (APS) is a rare autoimmune disease characterized by the occurrence of thrombotic events and obstetrical complications in the presence of antiphospholipid antibodies (aPL) [1]. The presence of aPL in conjunction with vascular damage can lead to other complications in APS patients, such as neurological, cardiac, and skin involvement [2]. APS is more frequent in women and is estimated to affect 50 per 100,000 population, with a mean diagnosis age of 50 years [3–5].

APS was originally described in patients with an underlying systemic autoimmune disease, commonly lupus (secondary APS) [6]. Nevertheless, in almost half of the cases, APS appears as an isolated disorder known as primary APS (PAPS) [7]. Beyond the presence of an underlying autoimmune disorder, no major clinical differences have been reported between the primary and secondary forms of APS [8].

Previous studies suggested the relevance of genetic susceptibility in primary and secondary APS [3, 8]. Several genetic loci within the human leukocyte antigen (HLA) class II region have been reported to be associated with PAPS, including *HLA-DRB1*1302* [3, 9]. Outside the HLA region, several loci associated with PAPS have been identified in candidate gene studies, including *STAT4*, *IRF5*, *MTOR*, and *PTPN22*. Despite these findings, genetic associations with PAPS outside of the HLA region have been inconsistent [3]. Further, no genome-wide association study (GWAS) has been previously performed in PAPS.

Herein, we perform a large GWAS and meta-analysis in PAPS including patients and controls from five different populations of European ancestry, and provide important new insights into the genetic basis and potentially the pathogenesis of PAPS.

## 2. MATERIALS AND METHODS

### 2.1. Study population and genotyping

Five independent case-control cohorts of European ancestry, with a total of 482 patients with PAPS and 5,003 controls, were studied. These include cohorts from Spain (90 PAPS-affected individuals and 1,517 unaffected controls), Italy (133 PAPS-affected individuals and 1,271 unaffected controls), Northern European and European-American (69 PAPS-affected individuals and 599 unaffected controls), European-American (90 PAPS-affected individuals and 1,047 unaffected controls) and an additional European cohort (100 PAPS-affected individuals and 569 unaffected controls). Genotyping data for the 1,047 control individuals included in the European-American cohort were derived from the database of Genotypes and Phenotypes (dbGaP, study accession: phs000187.v1.p1) [10]. All PAPS patients in this study met the Sydney classification criteria for primary APS [11]. Genomic DNA was extracted from blood samples by standard methods and genotyping data were generated via Illumina platforms (Illumina, San Diego, CA, USA) according to the manufacturer’s instructions (**Table S1**). The study was approved by the Ethics Committees at all participating institutions. Protocols followed the principles of the Declaration of Helsinki, and all individuals included in the study signed written informed consents.

### 2.2. Quality controls and imputation

Quality controls (QCs) of genotype data were performed for each cohort separately using Plink v.1.9 [12]. Individuals with a genotyping call rate <95% and those related or duplicated according to their identity by descent proportion (PI_HAT >0.4) were excluded from further analyses. Single-nucleotide polymorphisms (SNPs) with a genotyping call rate <98%, minor allele frequency (MAF) <1%, or those deviated from Hardy-Weinberg equilibrium (HWE) in cases and controls (p-value <1×10^−3^) were removed. In addition, A/T-C/G SNPs with a MAF close to 50% were removed from genotype data to avoid errors during the imputation. Filtered genotype data were imputed using the TOPMed Imputation Server with Minimac4 [13] and the TOPMed version R2 reference panel [14]. In the case of the European-American cohort, imputation of cases and controls was performed separately due to the low number of overlapping SNPs among the genotyping arrays. After imputation, SNPs with MAF <1%, HWE p-value<1 x 10^−3^, or imputation quality metric RSQ <0.9 were filtered out. A principal component analysis (PCA) was performed to control for population stratification using Eigensoft 6.1.4 [15] (**Figure S1**). This software calculates principal components in each population by using around 100,000 independent SNPs and estimating individuals’ standard deviations from cluster centroids. Thus, those individuals with >6 standard deviations were considered outliers and removed from further analyses. The genomic inflation factor (λ) was calculated for each cohort using the R package “gap”, and quantile-quantile (Q-Q) plots were generated. All plots were produced with R version 4.2.0.

Imputation of HLA classical alleles within the expanded HLA region (chr6: 20 to 40 Mb) was performed using the Michigan Imputation Server with Minimac4 [13], following the same procedure as for the genome-wide imputation in all the cohorts. HLA class I and II classical alleles were imputed for each cohort at 2 fields of resolution. Only classical alleles with an imputation quality metric RSQ >0.9 were used for further analyses.

### 2.3. Data analysis

Logistic regressions were conducted in each cohort by adjusting for the first five principal components using Plink v.1.9 software [12]. The summary-level statistics of each cohort were meta-analyzed using the inverse variance method. Thus, variants with no evidence of heterogeneity (Cochran’s Q test p-value >0.1 and heterogeneity index <50%) were considered under a fixed-effects model, while in variants that showed heterogeneity of effects between cohorts (Cochran’s Q test p-value ≤0.1 and heterogeneity index ≥50%) a random-effects model was applied. The genome-wide significance level was established at a p-value <5×10^−8^ and the suggestive association threshold was set at a p-value <1×10^−5^. To identify independent significant associations, we performed dependency analyses at the population level followed by inverse variance meta-analysis within and outside the HLA region. For this, the most associated variant identified in the meta-analysis for each significant locus at the genome-wide level was used as a covariate to perform the conditional logistic regression in each population. Then, the results from the conditional logistic regressions for each population were meta-analyzed, and the variant showing the most significant association was considered an independent association. Using a similar approach, a pairwise conditional analysis was performed within the HLA region to test the independence of the SNP identified in the meta-analysis and the classical allele associated with PAPS (p-value <1×10^−5^).

### 2.4. Functional annotation and enrichment analysis

*In silico* approaches were used to evaluate potential biological causality of significant and suggestive independent SNPs (linkage disequilibrium, LD r^2^<0.4). To assess their genomic location and possible functional implication the Open Targets Genetic [16, 17], VannoPortal [18], HaploReg v4.2 [19], Genotype-Tissue Expression (GTEx) project [20], and RegulomeDB [21, 22] databases, and the WashU Epigenome browser [23] were used. In order to assess enrichment in histone marks, the software GARFIELD was used to explore regulatory features from the Roadmap Epigenomics project [24]. Briefly, this function assesses functional annotation from GWAS summary statistics using several p-value thresholds (p-value ≤1×10^−5^, p-value ≤1×10^−4^, p-value ≤1×10^−3^, p-value ≤1×10^−2^) and considering LD, minor allele frequencies, and distance to the nearest gene of each variant.

Finally, key pathways were identified by a Gene Ontology (GO) enrichment analysis using the closest genes to significant and suggestive independent signals and using the Metascape web-based portal [25]. Those Biological Process ontology terms with p-value <1×10^−3^ and at least 3 genes in the GO term were reported.

### 2.5. Cumulative genetic risk score

To assess the variability of genetic risk to PAPS across populations, a cumulative genetic risk score (GRS) was calculated. For this analysis, a total of 2,504 individuals from the 1000 Genomes Project phase 3 major populations (African, n=661; Admixed American, n=347; East Asian, n=504; European, n=503; South Asian, n=489) were included [26]. Independent significant and suggestive variants at each locus reported in our meta-analysis were included in calculating the GRS, analyzing a total of 40 independent genetic variants (**Table S2**). For the variant located in the *ESR2* locus, a SNP in complete LD (r^2^ = 1) was used due to the lack of genetic information for the most significant SNP in this locus in the 1000 Genomes Project data. For GRS estimation, individuals’ genotypes were coded as 0, 1 or 2 indicating the number of PAPS risk alleles for each SNP. For each variant, the OR obtained from the meta-analysis was used. Thus, cumulative GRS was obtained by multiplying the natural logarithm of the OR for each SNP by the number of risk alleles for its corresponding SNP 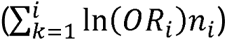 [27]. To determine whether the cumulative GRS was significantly different across populations, a Welch’s t-test was applied.

### 2.6. Relationship between PAPS genetic susceptibility with other immune-mediated diseases

We evaluated the relationship of PAPS with other immune-mediated diseases (IMDs) from a genetic perspective. A low-dimensional summary of the shared genetic component of selected IMDs, excluding the HLA region, was created using a PCA strategy to generate an object known as “basis” that comprises the multidimensional components of the shared genetic risk [28]. PAPS GWAS data were projected on this “basis” occupying a location with respect to the shared genetic architecture of the included IMDs. Based on this information, we estimated the closeness of PAPS to the rest of the IMDs by using a complete linkage hierarchical clustering method. However, the clustering algorithm can be unstable when considering multiple dimensions, so we also compared PAPS to IMDs’ projections by using the Mahalanobis distance. This distance was calculated as *(P_j_ – P_PAPS_)*′ *S^−1^ (P_j_ − P_PAPS_)* where *P_j_* and *P_PAPS_* represent the projected vectors of trait *j* and PAPS, respectively, and *S* represents the correlation derived from the LD among the SNPs in the principal components. When projections from GWAS data are used in clustering, the uncertainty associated with each observation can vary systematically with sample size, especially in studies with small sample sizes. To address this, we complemented the above clustering methods with the Dirichlet Process Mixtures with uncertainty (DPMUnc) approach [29], a Bayesian clustering method that takes the uncertainty associated with each data point into consideration.

With the purpose of identifying the genetic variation behind the shared signals between PAPS and other IMDs, we selected the SNPs that influence the projections of each significant PC in PAPS, referred to as driver SNPs. The probability of being significant in both PAPS and another IMD was estimated for each driver SNP using an FDR-based approach, considering a pairwise FDR <0.05 to be significant:

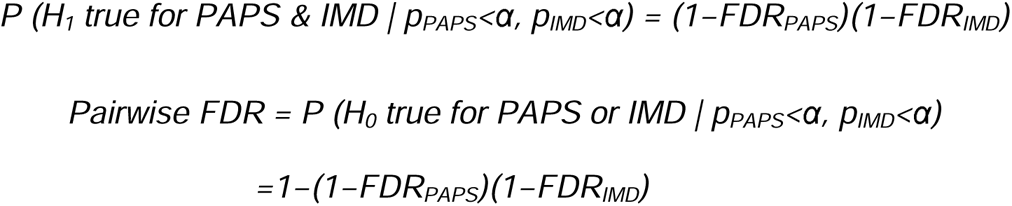

Driver SNPs represent a small fraction of the genome and they may be useful in the identification of potential true causal SNPs in LD with them for the diseases evaluated. In addition, true casual variants in LD with each driver SNP may differ between diseases. To address this, we used the colocalization method [30] to formally investigate whether a putatively causal variant is shared between PAPS and other IMDs within a specific locus. This method was applied on 2 Mb regions centered on each driver SNP with no overlapping among regions. Thus, driver SNPs and the variants within these regions were considered for colocalization. SNPs with the highest posterior probability of being the shared causal variant in each region were reported.

## 3. RESULTS

### 3.1. GWAS results and meta-analysis

Following genotype quality control measures and imputation, genetic data from a total of 5,222 individuals (410 patients with PAPS and 4,812 controls) were included in our genetic analysis (**Table S3**). A logistic regression adjusting for the first five PCs was performed on each independent cohort (**Figure S2**), followed by a meta-analysis of the summary statistics for all cohorts (**Figure 1**). After correction for population structure, deviation in genomic inflation factors was not detected in any cohort (**Figure S3 and Table S3**). The results of the meta-analysis revealed two associated loci (represented by 34 variants) at the genome-wide level of significance (p-value<5×10^−8^) and 349 suggestive associated variants (p-value<1×10^−5^) (**Table S4**). The significant associations were located within the *STAT4* and the HLA region (**Table 1**). The most significant variant in *STAT4* corresponds to an intronic SNP (rs11889341, p-value=1.39×10^−9^, OR [95% CI]=0.61 [0.52-0.71]; **Figure 2A**), showing a consistent effect in all five cohorts analyzed. Regarding the association within the HLA region, the most significant SNP is located near the transcription start site of the *HLA-DRA* gene (rs9269041, p-value=2.07×10^−8^, OR [95% CI]=0.63 [0.54-0.74]; **Figure 2B**).

**Figure 1.**
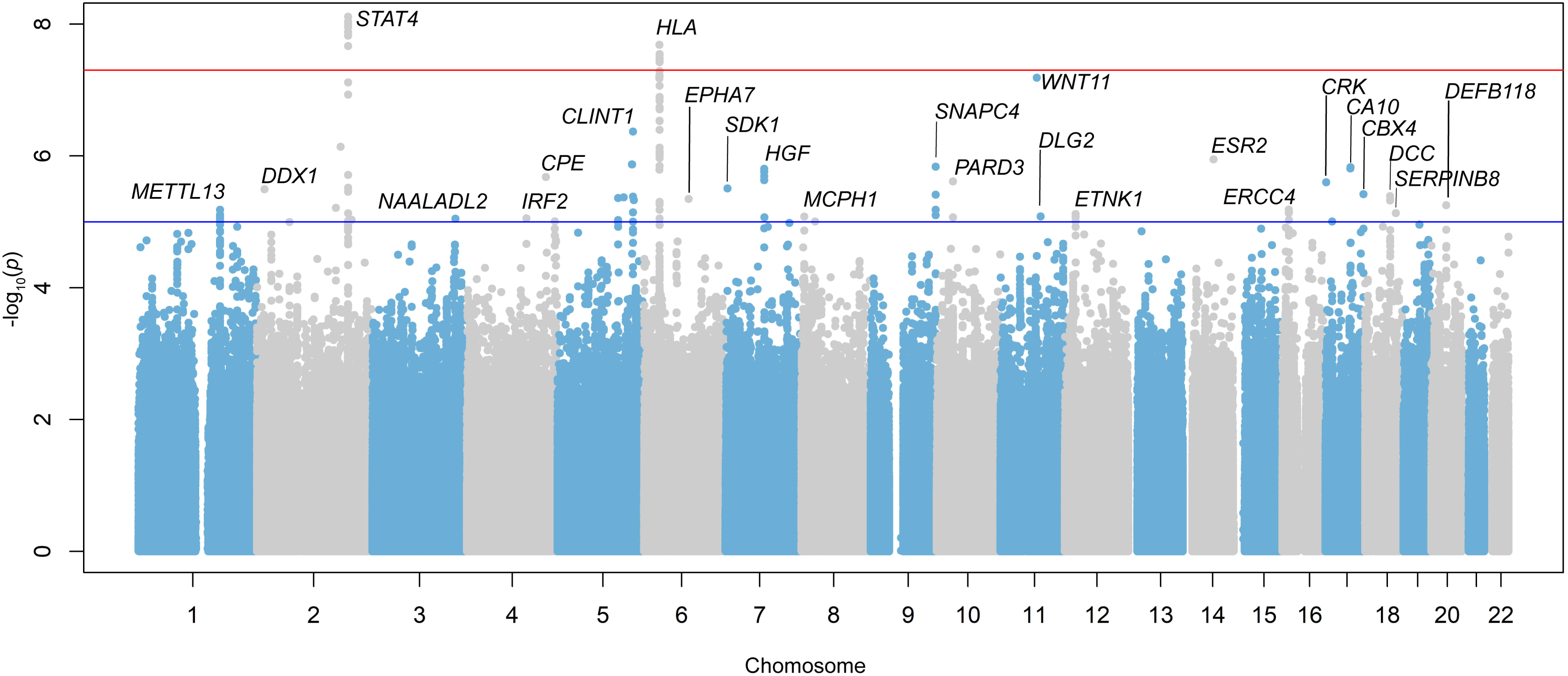
A Manhattan plot depicting the results of the meta-analysis among the five PAPS cohorts included in this study. Y and X axes refer to the –log_10_ p-values and chromosome positions, respectively. The red horizontal line represents the genome-wide association threshold (p-value <5×10^−8^) and the blue line represents the suggestive threshold (p-value<1×10^−5^).

**Figure 2.**
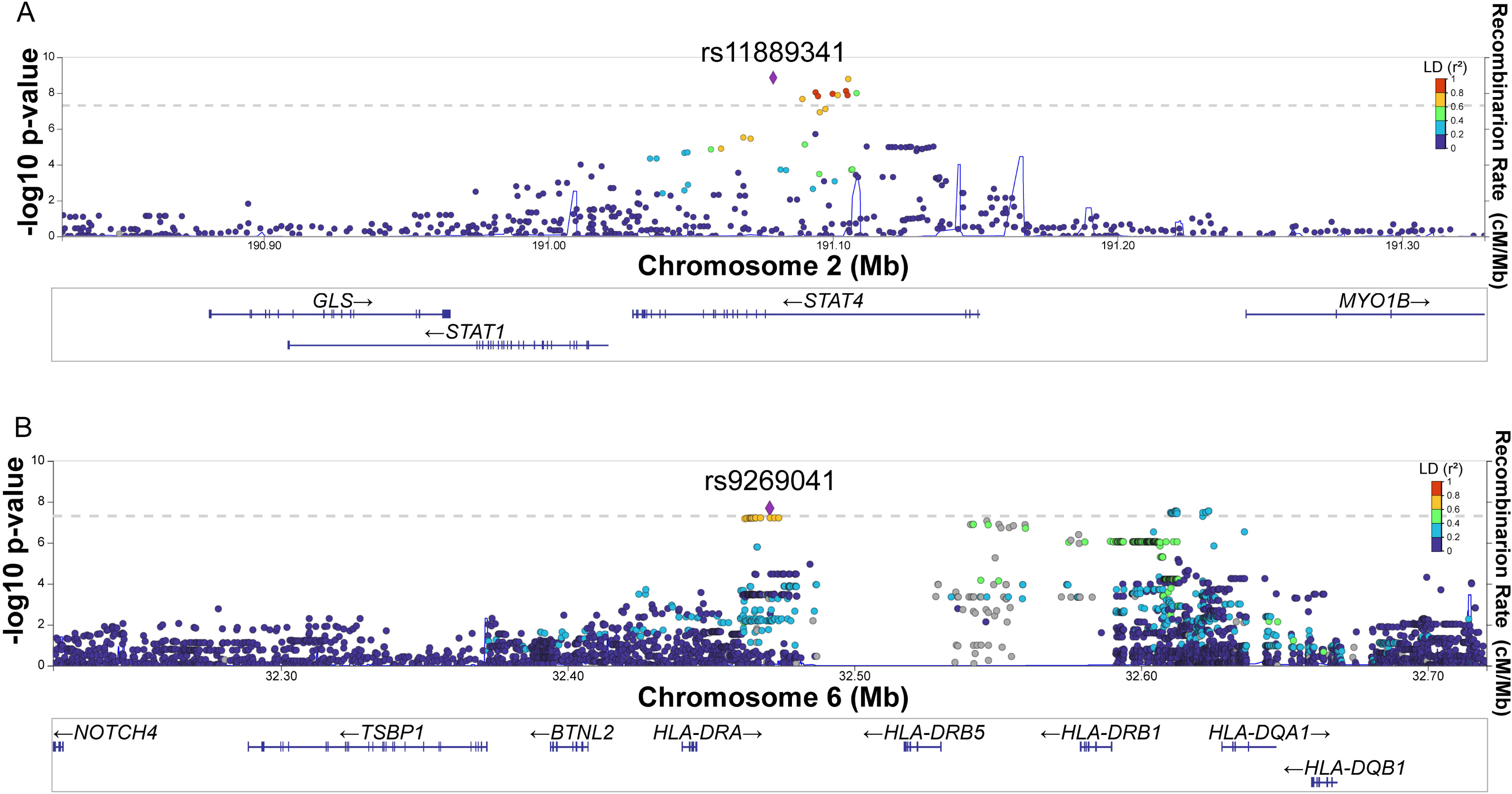
Regional plots for the genome-wide associated signals in chromosome 2. (**A**) and chromosome 6 (**B**), respectively. The y-axis corresponds to the –log_10_ p-values of evaluated variants while the x-axis indicates variants’ positions. Purple diamonds correspond to the most associated variants in each region. The rest of the variants are colored according to their degree of linkage disequilibrium (LD) with each top signal based on pairwise r^2^ values from the European population from the 1000 Genomes Project. The grey dashed line corresponds to the genome-wide association threshold (p-value<5×10^−8^) and the blue line to the estimated recombination rates. Genes within the selected regions are shown at the bottom of the plots.

**Table 1.** Meta-analysis results showing the most significant genetic variant in each locus associated with PAPS at a genome-wide level of significance (p-value<5×10^−8^)

| LOCUS | CHR | BP (HG38) | SNP | MINOR ALLELE | OR | 95% CI | P-VALUE |
| --- | --- | --- | --- | --- | --- | --- | --- |
| *STAT4* | 2 | 191079016 | rs11889341 | C | 0.61 | 0.52-0.71 | 1.39x10<sup>-9</sup> |
| *HLA-DRA* | 6 | 32470466 | rs9269041 | G | 0.63 | 0.54-0.74 | 2.07x10<sup>-8</sup> |

Abbreviations: Bp, base pair; chr chromosome; CI, confidence interval; OR, odds ratio; SNP, single-nucleotide polymorphism.

To assess independent significant associations with PAPS (p-value<5×10^−8^) in the *STAT4* and the HLA regions, we conducted stepwise conditional analyses in these two loci. As a result, the most significant SNPs from each locus, i.e. rs11889341 for *STAT4* and rs9269041 for the *HLA-DRA* region, explained the entire genetic association in these two regions (**Figure S4**).

Next, we imputed HLA classical alleles to determine their association with PAPS by using a specific reference panel for this region. *HLA-DRB1*1302* (p-value=1.12×10^−6^, OR=2.06) was the classical allele showing the most significant association with PAPS. The results of the logistic regressions and meta-analysis of HLA classical alleles after conditioning by *HLA-DRB1*1302* did not reveal any other suggestive associations (p-value<5×10^−5^). To determine if the signal detected in the *HLA-DRB1*1302* classical allele is independent of the PAPS-associated SNP close to *HLA-DRA,* i.e. rs9269041, we performed a pairwise conditional analysis. The results of this analysis showed a persistent, albeit slightly attenuated, genetic effect in each signal after adjusting for the other, suggesting an independent effect of these two markers in PAPS (**Table 2**).

**Table 2.** Pairwise conditional genetic analysis between rs9269041 and *HLA-DRB1*13:02* in PAPS compared to healthy controls.

| MARKER | COVARIATES | P-VALUE | OR |
| --- | --- | --- | --- |
| rs9269041 | None | $2.07 \times 10^{-8}$ | 0.63 |
| | *HLA-DRB1\*13:02* | $9.10 \times 10^{-7}$ | 0.66 |
| *HLA-DRB1\*13:02* | None | $1.12 \times 10^{-6}$ | 2.06 |
| | rs9269041 | $1.76 \times 10^{-4}$ | 1.76 |

Abbreviations: OR, odds ratio.

In addition to the genetic variants showing association with a genome-wide level of significance, a total of 349 SNPs in the meta-analysis surpassed the suggestive significance threshold (p-value<1×10^−5^) (**Table S4**). Including these, a total of 40 SNPs correspond to independent variants (r^2^<0.4) located within 36 loci that are associated with PAPS with either a genome-wide level or suggestive level of significance (**Table S5**). Notably, some of these suggestive loci have been associated with clinical traits or molecular pathways that might be manifested in patients with PAPS. *ESR2* (estrogen receptor 2) (rs932657402, p-value=1.13×10^−6^, OR [95% CI]=0.13 [0.06-0.30]) and *HGF* (hepatocyte growth factor) (rs75427037, p-value=1.57 x 10^−6^, OR [95% CI]=0.23 [0.12-0.42]), have been associated with thrombotic events [31, 32], while *EPHA7* (rs117489460, p-value=4.50 x 10^−6^, OR [95% CI]=0.19 [0.09-0.38]) is expressed in vascular endothelial cells and shows higher expression levels during inflammation [33]. Other suggestive loci identified in our meta-analysis have been previously associated with other immune-mediated diseases, such as *WNT11* (Wnt family member 11) and *DLG2* (discs large MAGUK scaffold protein 2) with inflammatory bowel disease, *ANKRD50* (ankyrin repeat domain containing 50) with systemic lupus erythematosus, and *PLCL1* (phospholipase C like 1) gene expression with rheumatoid arthritis [34–37].

Logistic regression for each cohort also revealed several loci with suggestive evidence of association with PAPS (p-value<1×10^−5^) (**Figure S2**).

### 3.2. Functional annotations and epigenetic enrichment analysis

In line with other autoimmune diseases, most PAPS-associated variants are in non-coding genetic regions and may influence the disease through regulatory elements. Therefore, we performed a functional annotation of significant and suggestive independent PAPS-associated variants (**Table S5**) by using *in silico* functional approaches.

First, we assessed the regulatory potential of significant and suggestive disease-associated independent variants identified in our meta-analysis. The functional annotation of these SNPs indicated their co-localization with epigenetic marks, suggesting a potential contribution to chromatin states across multiple tissues and cell types (**Table S6**). We found that 37 of these 40 disease-associated SNPs may alter regulatory motifs, which might affect transcription factor (TF) binding. DNase hypersensitivity sites were observed in eight of these variants, three of them also in protein binding sites. When all the significant and suggestive variants from the meta-analysis were evaluated for enrichment in histone marks, three histone modifications showed a significant enrichment. These marks corresponded to the H3K4me3 (p-value=3.62×10^−2^), H3K4me2 (p-value=1.12×10^−2^), and H4K20me1 (p-value=3.05×10^−2^) in blood cells, specifically in GM12878 lymphoblastoid cells.

Given the effect on gene expression that epigenetic marks can exert, we evaluated the relationship of significant and suggestive independent variants located at these epigenetic marks with changes in gene expression by examining eQTL. A total of eight PAPS-associated variants were linked to gene expression variation of at least one gene in multiple tissues (**Table S7**). The most significant eQTLs were identified for the variant associated with PAPS near *HLA-DRA*. The disease risk in this SNP (rs9269041) was associated with increased expression of several HLA class II genes, including *HLA-DRB6*, *HLA-DRB9*, *HLA-DPB2*, *HLA-DQA2* and *HLA-DQB2* in whole blood, vascular tissue, and nervous tissue (**Figure 3, and Figure S5**). Experimental ChIP-seq publicly available data show an epigenetic mark of active enhancer (H3K27ac) near rs9269041 in CD14+ monocytes, further supporting a regulatory potential for this PAPS-associated genetic susceptibility locus (**Figure S6**).

**Figure 3.**
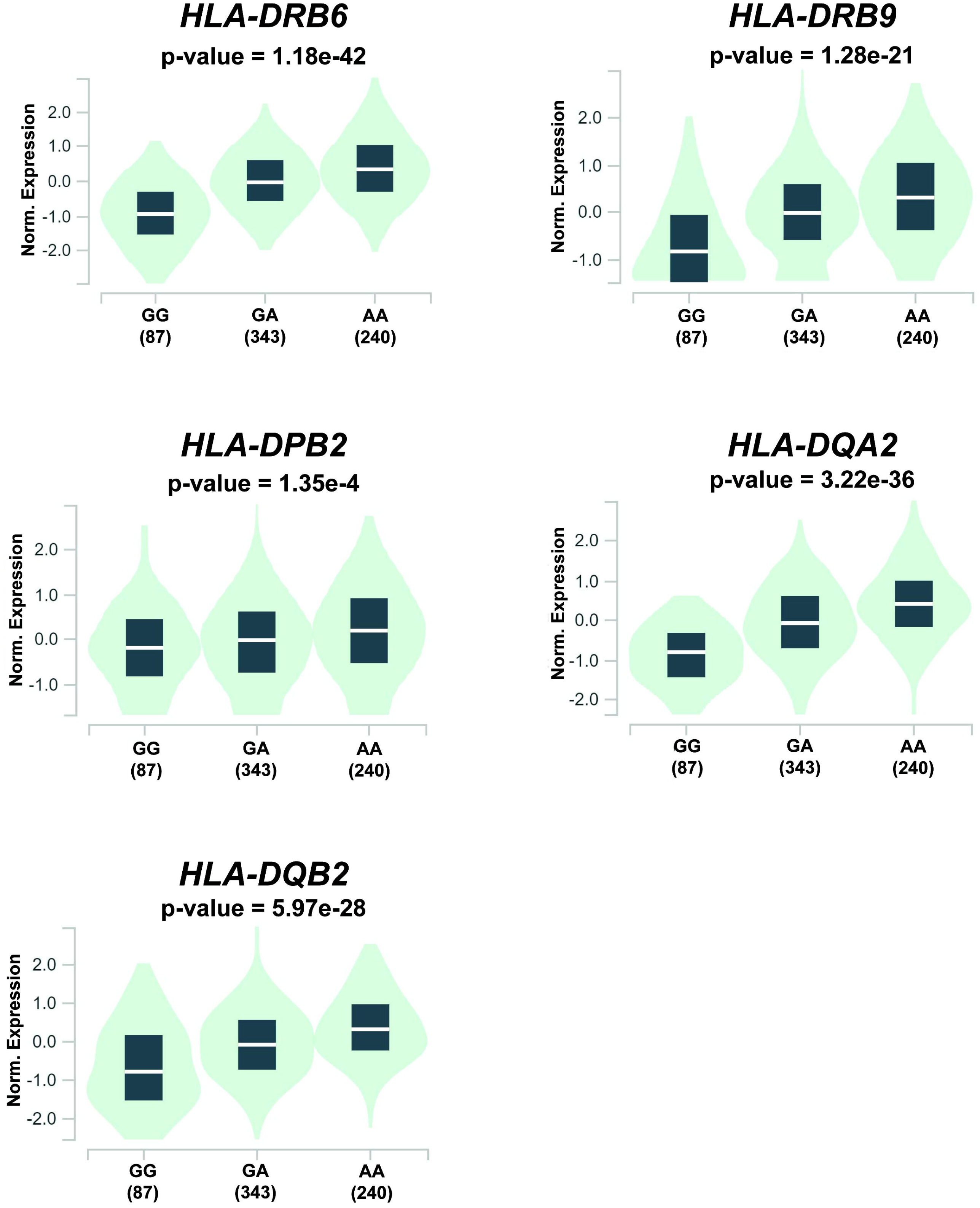
Violin plots representing the differences in gene expression levels in whole blood (y-axis) of the HLA genes *HLA-DRB6*, *HLA-DRB9*, *HLA-DPB2*, *HLA-DQA2* and *HLA-DQB2* depending on the genotypes of the PAPS-associated variant within the HLA region, rs9269041 (x-axis). When the PAPS-risk allele is present (rs9269041-A), higher gene expression levels are observed.

Furthermore, two PAPS-associated SNPs alter the gene expression of their nearest gene, producing expression changes in *SGCD* and *METTL13* in several tissues, including cardiovascular system tissues and the skin (**Table S7**).

Chromatin conformation might alter transcriptional regulation through physical interactions with other regions of the genome. To examine the presence of physical interactions within the regions containing PAPS-associated variants with gene bodies and/or gene promoters, the VannoPortal and the Open Targets Genetics public databases were used. From the list of significant and suggestive independent PAPS-associated variants, we identified 25 SNPs establishing physical interactions with gene bodies in the GM12878 lymphoblastoid cell line, and 11 promoter interactions in several other cell types (**Table S8**). The majority of these variants interact with multiple genes, some of them with genes whose expression levels were affected (**Table S9**). Some of these interactions are noteworthy given their functional relevance, as they are supported by eQTL evidence of increased gene expression in the presence of the risk alleles. These interactions correspond to the PAPS-associated variants rs35930163 and rs9411271 with the *DNM3* and *GPSM1* genes, respectively (**Figure 4**). These functional annotations suggest these loci as potential target genes for PAPS.

**Figure 4.**
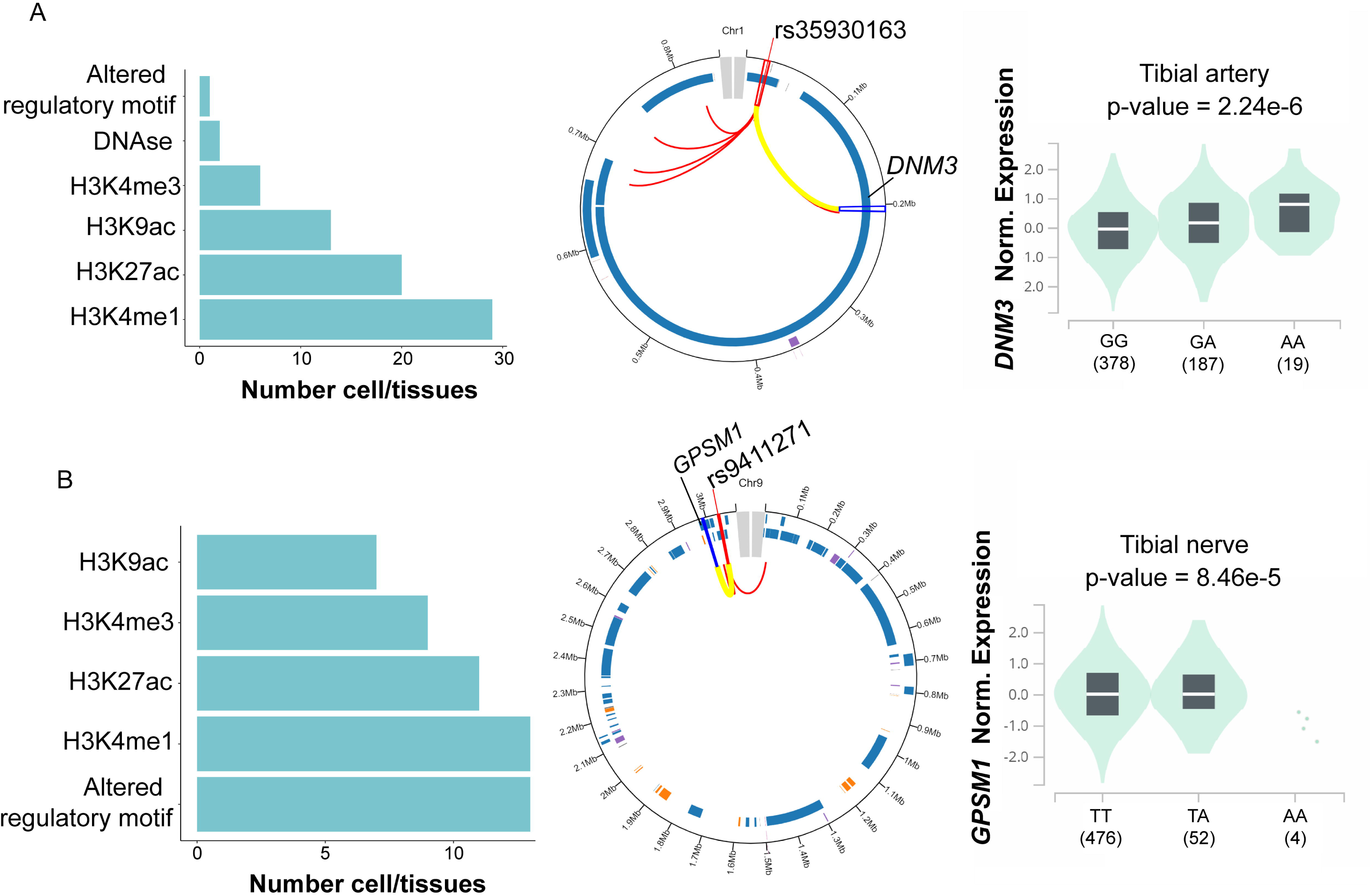
Functional annotation results for the variants. (**A**) rs35930163 and (**B**) rs9411271 located close to the genes *DNM3* and *GPSM1*, respectively. Left panels indicate epigenetic mark binding at each variant. Central panels represent the physical chromatin interactions established by the regions containing each variant in the lymphoblastoid cell line GM12878; the red lines represent all the interactions of each region while the yellow ones refer to the interaction with the indicated genes. The right panels show violin plots representing the differences in expression levels (y-axis) of the interacting genes depending on the genotypes of each PAPS-associated variant (x-axis).

### 3.4. Pathway analysis

To identify possible biological processes enriched in the loci identified in the meta-analysis, we performed a GO term enrichment analysis including the closest genes to the significant (p-value<5×10^−8^) and suggestive (p-value<1×10^−5^) independent variants associated with PAPS. Six different clusters of GO terms were identified to be enriched in the reported loci: establishment or maintenance of cell polarity (p-value=1.42×10^−7^), cell junction assembly (p-value=8.73×10^−7^), negative regulation of cell growth (p-value=7.39×10^−5^), establishment of cell polarity (p-value=5.31×10^−4^), regulation of hydrolase activity (p-value=2.12×10^−3^), and regulation of leukocyte mediated immunity (p-value=3.20×10^−3^). Interestingly, PAPS-associated genes showed enrichment in biological processes related to the central nervous and the immune systems (**Table S10)**.

### 3.5. Cumulative GRS

Significant and suggestive independent variants identified in the meta-analysis were used to evaluate the genetic risk for PAPS in 2,504 individuals from the five major populations included in the 1000 Genomes Project. The highest genetic risk score (GRS) for PAPS was observed in the East Asian and African populations, while the lowest genetic risk was detected in the European and South Asian populations (**Figure 5**). When mean GRS values were compared, all populations showed significant differences in the pairwise comparisons except for the Admixed American versus South Asian populations (Welch’s t-test p-value=0.25).

**Figure 5.**
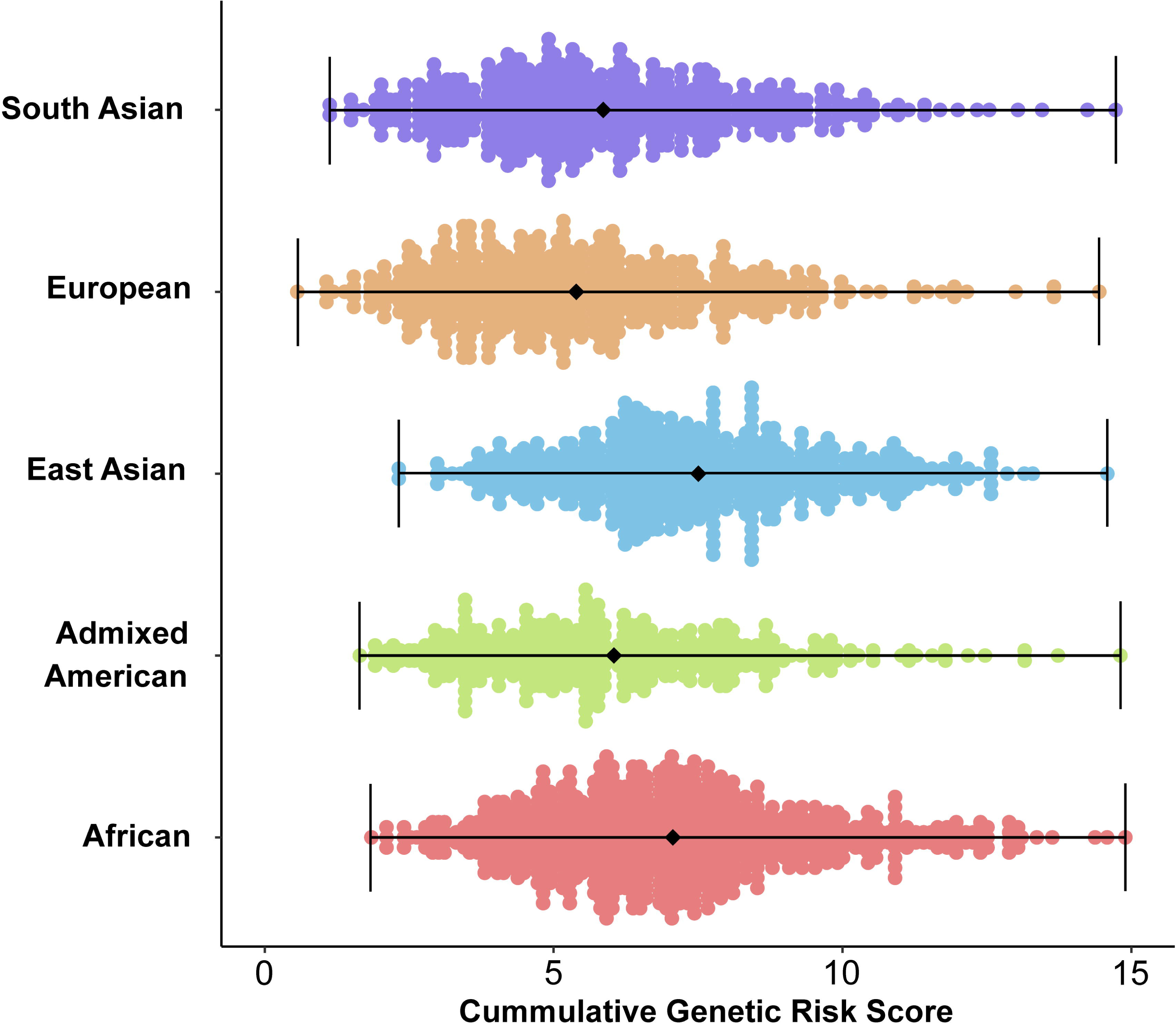
Dot plot of the cumulative genetic risk score (GRS) for PAPS susceptibility across the major populations of the 1000 Genomes project.

### 3.6. Relationship of PAPS with other immune-mediated diseases

The evaluation of the common genetic variation among traits or diseases can be useful in the identification of shared pathways that may be involved in the pathophysiology of a disease. To assess whether PAPS susceptibility variants might be shared with other traits or diseases, we conducted a search for significant and suggestive independent variants identified in our meta-analysis in public PheWAS data. From the 40 independent SNPs associated with PAPS in the meta-analysis, 14 showed a significant association (p-value<0.05) in PheWAS data with immune-related traits (**Table S11**). The most significant associations were observed for SNPs located in the *HLA-DRA* and *STAT4* loci, while the diseases showing a higher number of associations with PAPS susceptibility loci were systemic lupus erythematosus and rheumatoid arthritis.

Comparing the results of genetic association studies has proven to be a useful tool in the identification of shared genetic variation among different diseases. To assess genetic similarities among PAPS and other IMDs, we projected PAPS summary statistics into a low-dimensional representation of the shared risk of selected IMDs distributed in 13 components [28]. We found PAPS differed significantly (FDR<5%) from controls on three of the 13 components, corresponding to the autoimmune (PC1), systemic lupus erythematosus (PC2), and the eosinophilic (PC13) signals (**Figure 6A**). The hierarchical clustering of the projections showed that PAPS is closer to IgG+ neuromyelitis optica (NMO) (**Figure 6A**), an inflammatory condition that affects the central nervous system [38]. This result was confirmed by the Mahalanobis and DPMUnc distance methods, which also suggested a close relationship of PAPS genetics with Sjögren’s syndrome, systemic sclerosis (SSc), juvenile idiopathic arthritis (JIA), CREST syndrome, and primary biliary cholangitis (PBC) (**Figure 6B** **and** **Figure 6C**).

**Figure 6.**
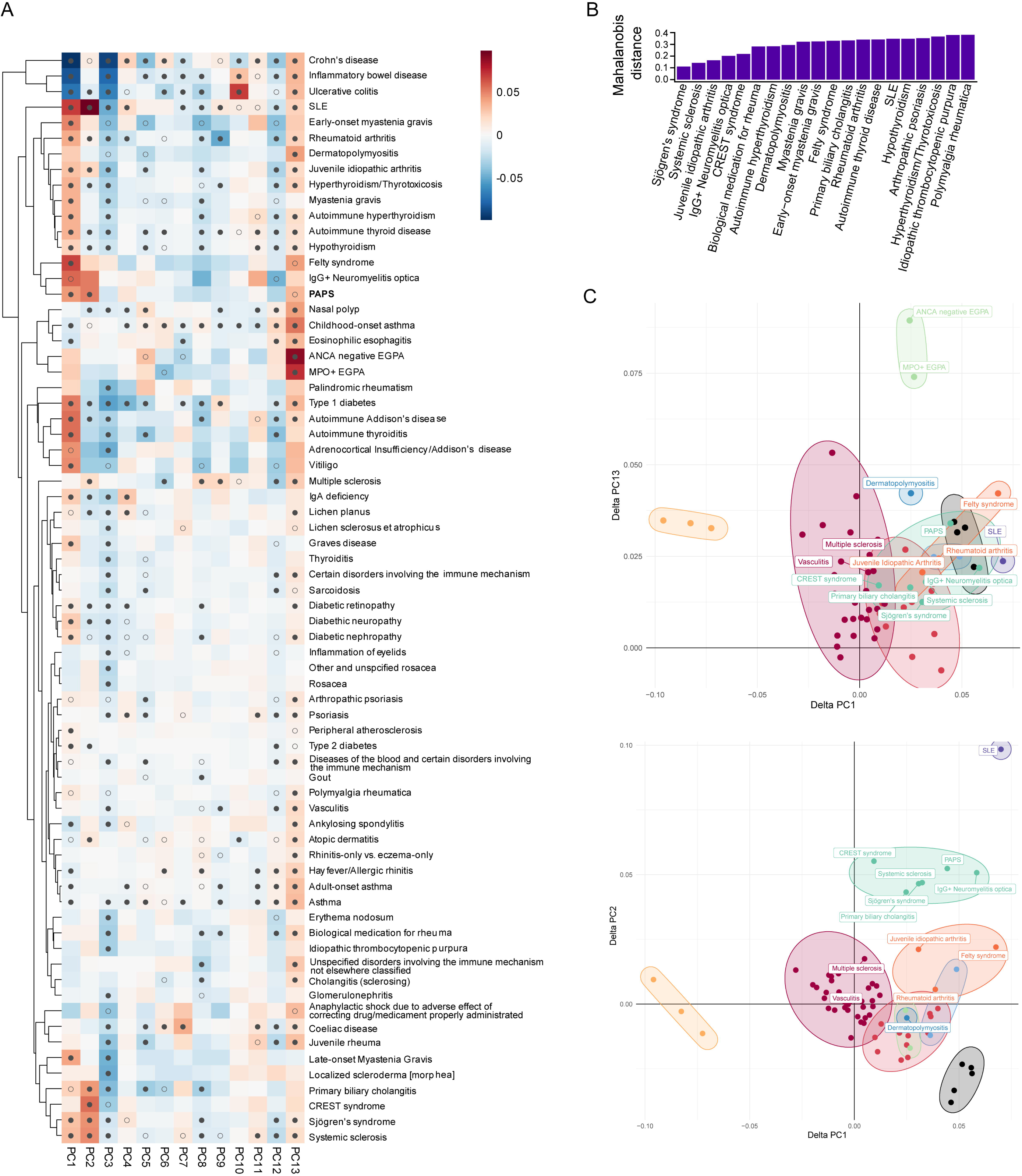
Results of etiologic similarity analyses of PAPS with other immune-mediated diseases (IMDs). (**A**) Hierarchical clustering of PAPS with other immune-mediated diseases GWAS data and self-reported disease traits in UK Biobank, FinnGen Project, and The International Multiple Sclerosis Genetics Consortium. PAPS appears in boldface. Heatmaps indicate delta values for each disease on each component PC1-PC13, with grey indicating 0 (no difference from control), and darker shades of blue or magenta showing departure from controls in one direction or the other. A filled circle indicates that the delta value was significantly non-zero with a significance of FDR <1%, while an unfilled circle indicates a significance of FDR <5%. (**B**) Top 20 closest Mahalanobis distances to PAPS of all tested traits. The Y axis displays the distance between the center of the distribution (PAPS) and different points (the IMDs). (**C**) Clustering analysis of PAPS with IMDs using the Dirichlet Process Mixtures with uncertainty method (DPMUnc). Features in which PAPS was significant at FDR 5% level are depicted, comparing PC1 versus PC13 (top) and PC1 versus PC2 (bottom).

Next, we assessed the variation underlying this shared genetic component. For this, 229 driver SNPs that influence the projections for which PAPS was significant, i.e., PC1, PC2, and PC13 signals, were selected. The probability of driver SNPs to be significant in PAPS-IMD pairs was estimated, selecting for this analysis those IMDs resulting from distance methods: CREST syndrome, SSc, Sjögren’s syndrome, IgG+ NMO, PBC, JIA, dermato/polymyositis, Felty syndrome, and localized scleroderma (morphea). We identified 9 significant PAPS-IMDs pairs comprising 4 driver SNPs that are significant in PAPS and other diseases from the pairs (**Table S12**). To assess whether these driver SNPs are acting as causal variants in both diseases of each PAPS-IMD pair or are close to causal variants, we applied a colocalization approach [30] by considering the genetic variation within a 1 Mb window for each driver SNP. Since driver SNPs on chromosome 7 (**Table S12**) are less than 1 Mb distant, only the one with the lowest pairwise FDR was considered for colocalization analysis. The colocalization analysis identified the most significant association in the *STAT4* locus from the meta-analysis as a causal variant in this region that is shared with JIA, PBC, and Sjögren’s syndrome (**Table S13**). Other SNPs in *TNPO3* and close to *BLK* were also identified as shared causal variants shared with SSc (**Table S13**).

## 4. DISCUSSION

We report the results of a large genetic association study in PAPS including affected and unaffected individuals of European ancestry. Two associations with a GWAS level of significance were identified near *HLA-DRA* and *STAT4*, contributing to PAPS genetic susceptibility. Further, 34 additional genetic susceptibility loci were identified with a suggestive level of association with the disease.

The most significant genetic association identified in our meta-analysis corresponds to a variant (rs11889341) located in *STAT4*, with a consistent effect in the five cohorts analyzed. *STAT4* encodes a signal transducer and activator of transcription and has been previously associated with PAPS in a candidate gene study [7]. In addition to PAPS, this variant has also been associated with other autoimmune conditions, including systemic lupus erythematosus and rheumatoid arthritis, and displaying a risk effect consistent with the results for PAPS [39]. The *STAT4* variant rs7574865, which is more extensively studied in systemic lupus erythematosus and rheumatoid arthritis, is in high LD (r^2^=0.98) with the PAPS-associated variant rs11889341. Indeed, rs7574865 also showed a significant risk association with PAPS in our study (p-value=1.11×10^−8^, OR=1.70).

We identified a genetic association between PAPS and the HLA class II region, which we localized to a genetic region near *HLA-DRA*. Transcription factor binding and eQTL analyses suggest that this effect alters the binding of multiple transcription regulators and the expression of several genes within the HLA region. The PAPS-associated risk allele in rs9269041, which tags the effect in *HLA-DRA*, is associated with overexpression of *HLA-DRB6*, *HLA-DRB9*, *HLA-DPB2*, *HLA-DQA2*, and *HLA-DQB2* in several tissues including whole blood and tissues relevant to PAPS pathogenesis such as the vascular and nervous systems. In addition, we identified an association between *HLA-DRB1*1302* and PAPS, which appears to be independent of the association in the *HLA-DRA* locus. The association between *HLA-DRB1*1302* and APS in cohorts combining both primary and secondary APS patients has been previously described [3]. Our study replicates and confirms this genetic association in the primary form of the disease.

Our study suggested a genetic susceptibility locus in *ESR2*, which encodes estrogen receptor 2, in patients with PAPS. Polymorphisms in this gene have been previously associated with the risk of deep vein thrombosis [31, 40]. Our data also suggested the genetic association between *HGF* and PAPS. *HGF* encodes hepatocyte growth factor and has been previously reported to be increased in the plasma in thrombosis-associated disorders in human and murine models [32, 41].

eQTL annotation revealed that the risk alleles of two PAPS-associated genetic variants, rs35930163 and rs9411271, increase the gene expression levels of *DNM3* and *GPSM1* genes. Indeed, chromatin interactions were revealed between these two disease-associated variants and gene body regions of *DNM3* and *GPSM1*. *DNM3* encodes Dynamin 3, which is a member of a family of binding proteins that have been previously associated with coagulation effects and venous thromboembolism in the Finngen public data [42]. *GPSM1* encodes the G Protein Signaling Modulator 1 that has been found to be related to alterations in cardiovascular function and regulatory mechanisms of the immune function in mouse models [43, 44].

We identified several biological processes enriched in PAPS-associated genetic susceptibility loci. The most significant gene ontologies correspond to the maintenance of cell polarity, cell junction organization, and cell adhesion processes. Previous studies have shown increased production of cell adhesion molecules in endothelial cells stimulated with antiphospholipid antibodies, which has been related to increased inflammation and thrombosis due to leukocyte binding to vascular walls [45, 46]. Interestingly, our enrichment analysis also highlighted leukocyte-mediated immunity, confirming the relevance of PAPS genetics in the immune system pathways. It is noteworthy that biological processes related to the regulation of the nervous system development were enriched in PAPS-associated loci. A high percentage of PAPS patients develop cognitive impairment, which in some cases precedes the diagnosis of the disease [47].

The cumulative GRS for PAPS suggested significant differences among populations. Individuals from East Asia, followed by African and Admixed American populations, showed the highest GRS values. The clinical variability in PAPS makes it challenging to accurately estimate the incidence and prevalence of this disease in different populations [4]. Despite this, population-based studies report a higher prevalence of PAPS, as well as a higher mortality, in Asian populations [48]. It is noteworthy that possible variability in the strength of genetic associations at the detected loci between populations might affect the accuracy of interpreting these results. Therefore, further validation studies in different populations and ethnic groups will be necessary.

Our genetic similarity analysis comparing PAPS to other immune-mediated diseases revealed a genetic relatedness for PAPS with NMO and Sjögren’s syndrome, both characterized by the development of neurological manifestations [49, 50]. Although the neurological involvement of PAPS is poorly understood, it has been primarily attributed to the thrombotic aspect of the disease [51]. A role for anti-NMDA antibodies in cognitive impairment in PAPS has also been suggested [52]. Our findings suggest that neurological manifestations in PAPS may be beyond the thrombotic aspect, reinforcing the concept of immune-mediated etiology in PAPS manifestations. In addition, non-thrombotic neurological manifestations have been described in APS [47]. The genetic similarity analysis also showed a relatedness between PAPS with SSc, specifically with limited cutaneous SSc (CREST), where endothelial cell dysfunction and vascular involvement are prominent [53, 54]. Indeed, endothelial dysfunction appears to play a role in the pathogenesis of PAPS [46, 55].

In summary, we performed a multi-center genome-wide association study in PAPS, and characterized genetic susceptibility loci associated with this rare disease. Our findings revealed novel genetic insights and identified new candidate genes associated with the pathogenies of PAPS. Genetic similarity analysis suggested relatedness between PAPS and other immune-mediated diseases characterized by neurological and vascular involvement. Additional studies with larger sample sizes and diverse populations are needed to confirm, replicate, and extend our results. Subsequent studies with adequate power to examine genetic susceptibility to specific clinical subphenotypes and serological specificities within PAPS are also warranted.

## Contributorship

– All authors fulfilled the following criteria:

– Substantial contributions to the conception or design of the work, or the acquisition, analysis or interpretation of data.
– Drafting the work or revising it critically for important intellectual content.
– Final approval of the version published.

## Supporting information

PRECISESADS Consortium

## Data Availability

All data are included in the manuscript or supplementary material.

## Acknowledgements

The authors would like to thank participating members of PRECISESADS for their contributions to this work (Supplemental information). This work was supported by the use of study data downloaded from the dbGaP website, under dbGaP: phs000187.v1.p1. The authors also acknowledge the participants and investigators of the FinnGen study.

## Ethical approval information

The study was approved by the Institutional Review Board of the University of Pittsburgh, Pittsburgh, Pennsylvania, USA.

## Data sharing statement

All data are included in the manuscript or supplementary material.

## Patient and public involvement

It was not appropriate or possible to involve patients or the public in the design, or conduct, or reporting, or dissemination plans of our research.

