## Supplementary Figures for "A genome-wide association study suggests new susceptibility loci for primary antiphospholipid syndrome"

**SUPPLEMMENTARY DATA**

**Figure S1.** Principal component (PC) analysis illustrating PC1 and PC2 in each individual population: Italian (a), Northern European and European-American (b), European (c), European-American (d) and Spanish (e). Cases and controls are shown as yellow and blue circles, respectively.

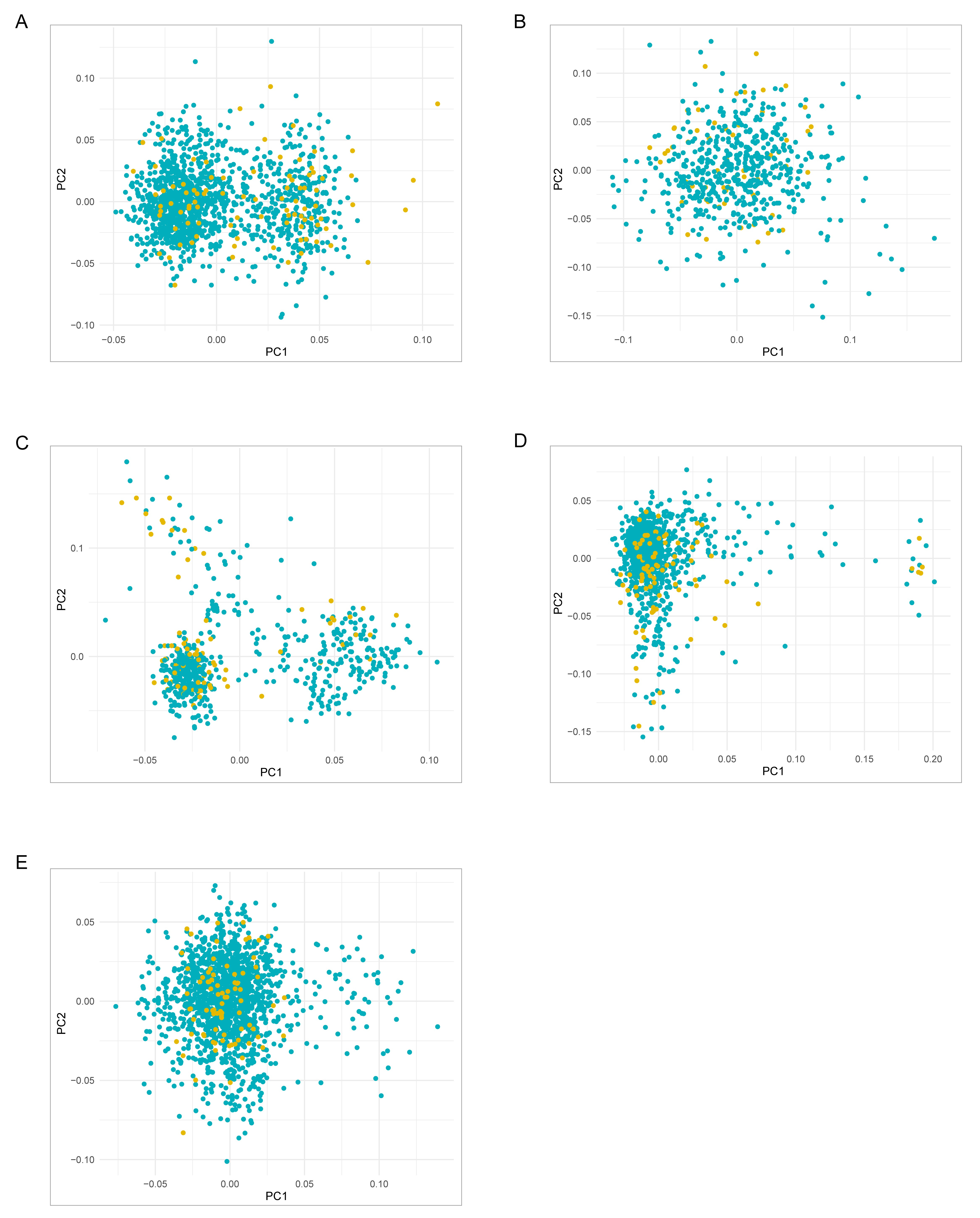

**Figure S2.** Manhattan plots depicting the GWAS results of each individual population: Italian (a), Northern European and European-American (b), European (c), European-American (d) and Spanish (e). The -log_10_ p-values for genetic variants analyzed are plotted against their physical chromosomal positions. The red and blue lines represent the genome-wide level of significance (p value<5x10^-8^) and the suggestive level of significance (p-value<5x10^-5^), respectively.

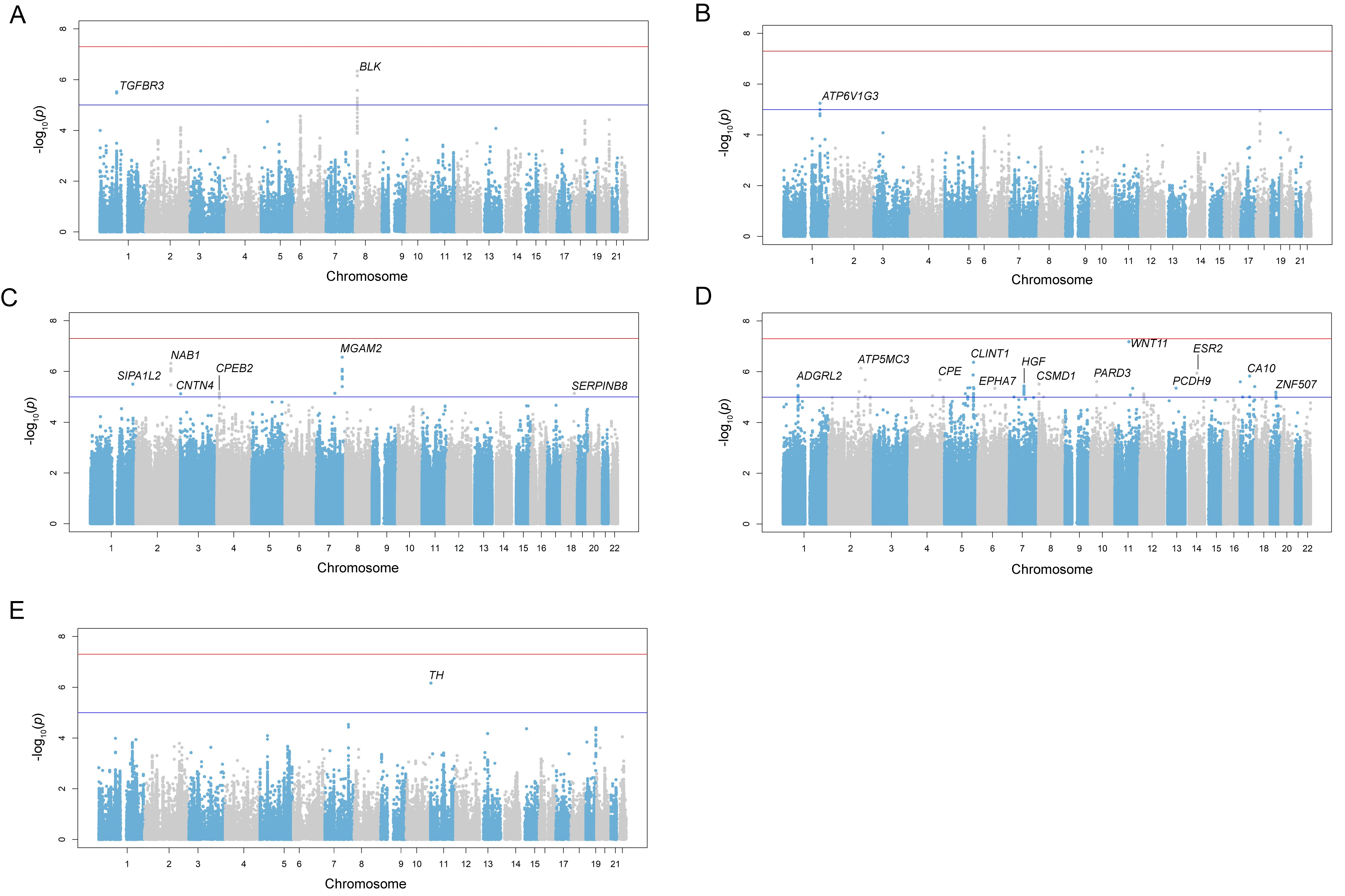

**Figure S3.** Quantile–quantile plots for the p-values of each individual population: Italian (a), Northern European and European-American (b), European (c), European-American (d) and Spanish (e). The x-axis indicates the expected distribution of -log_10_ (p-values) and the y-axis indicates the observed distribution of -log_10_ (p-values). Genomic inflation of the observed distribution is indicated by λ.

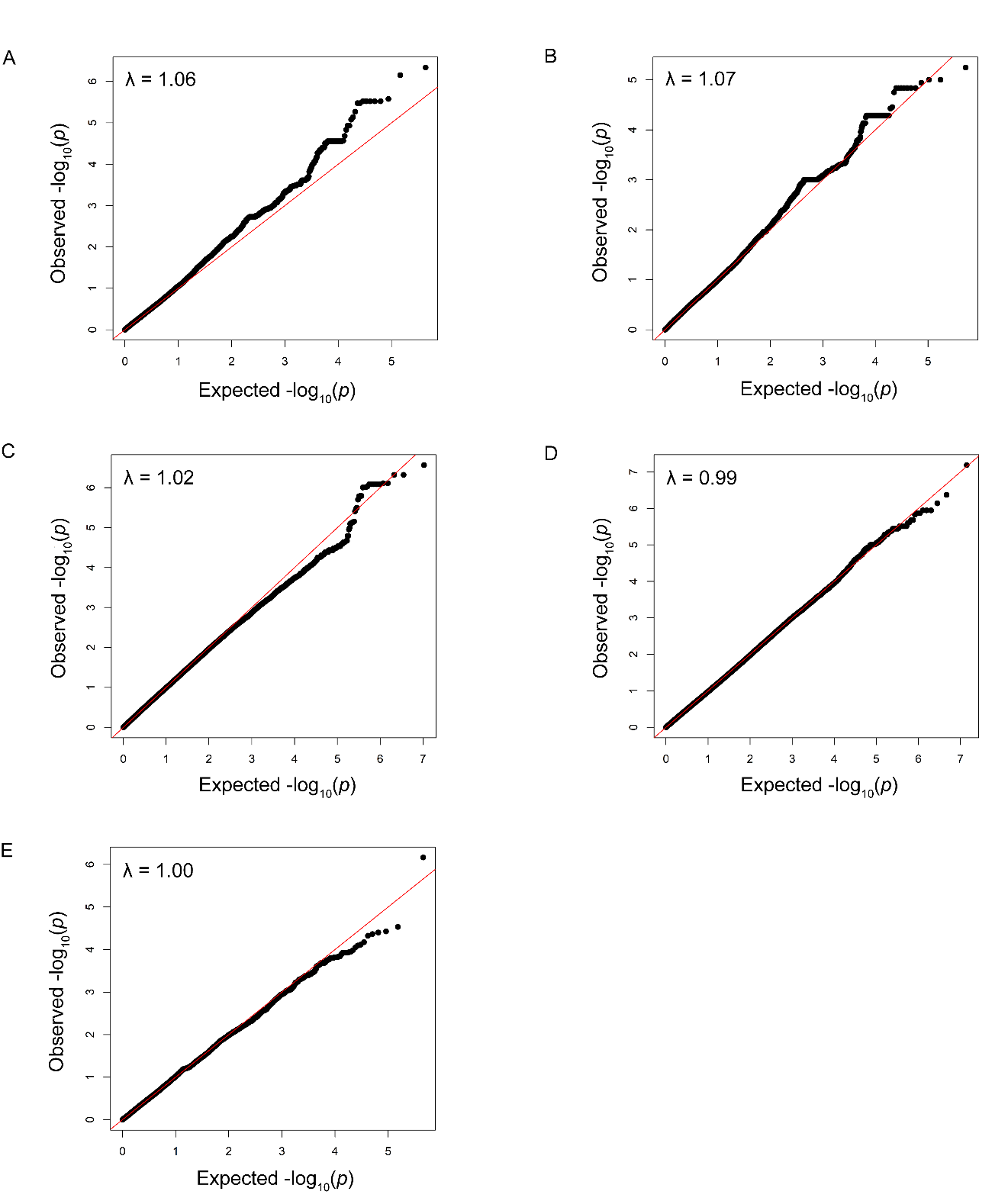

**Figure S4.** Regional Manhattan plots showing the results of stepwise conditional analyses in the *STAT4* and HLA regions from the genome-wide meta-analysis. (A) Unconditioned in *STAT4* region; (B) conditioning on rs11889341 in *STAT4* region; (C) unconditioned in HLA region; (D) conditioning on rs9269041 in HLA region. The -log_10_ p-values for each genetic variant analyzed are plotted against their physical chromosomal positions. The red and blue lines represent the genome-wide level of significance (p-value<5x10^-8^) and the suggestive level of significance (p-value<5x10^-5^), respectively.

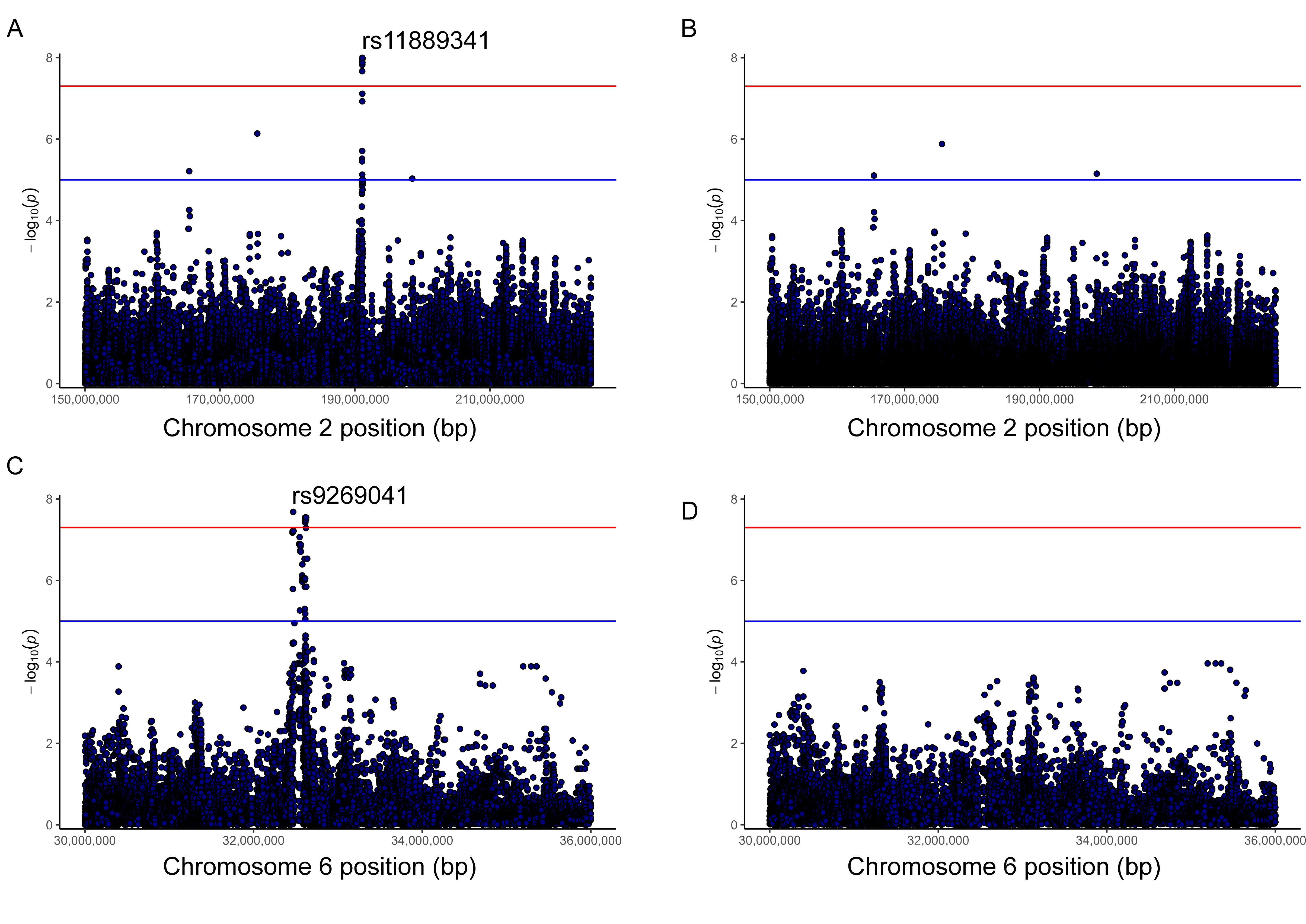

**Figure S5.** Violin plots representing the differences in expression levels (y-axis) of the HLA genes *HLA-DRB6*, *HLA-DRB9*, *HLA-DPB2*, and *HLA-DQB2* depending on the genotypes of the PAPS-associated variant rs9269041 within the HLA region (x-axis). When the PAPS-risk allele is present (rs9269041-A), higher gene expression levels are observed in coronary artery and tibial nerve tissues.

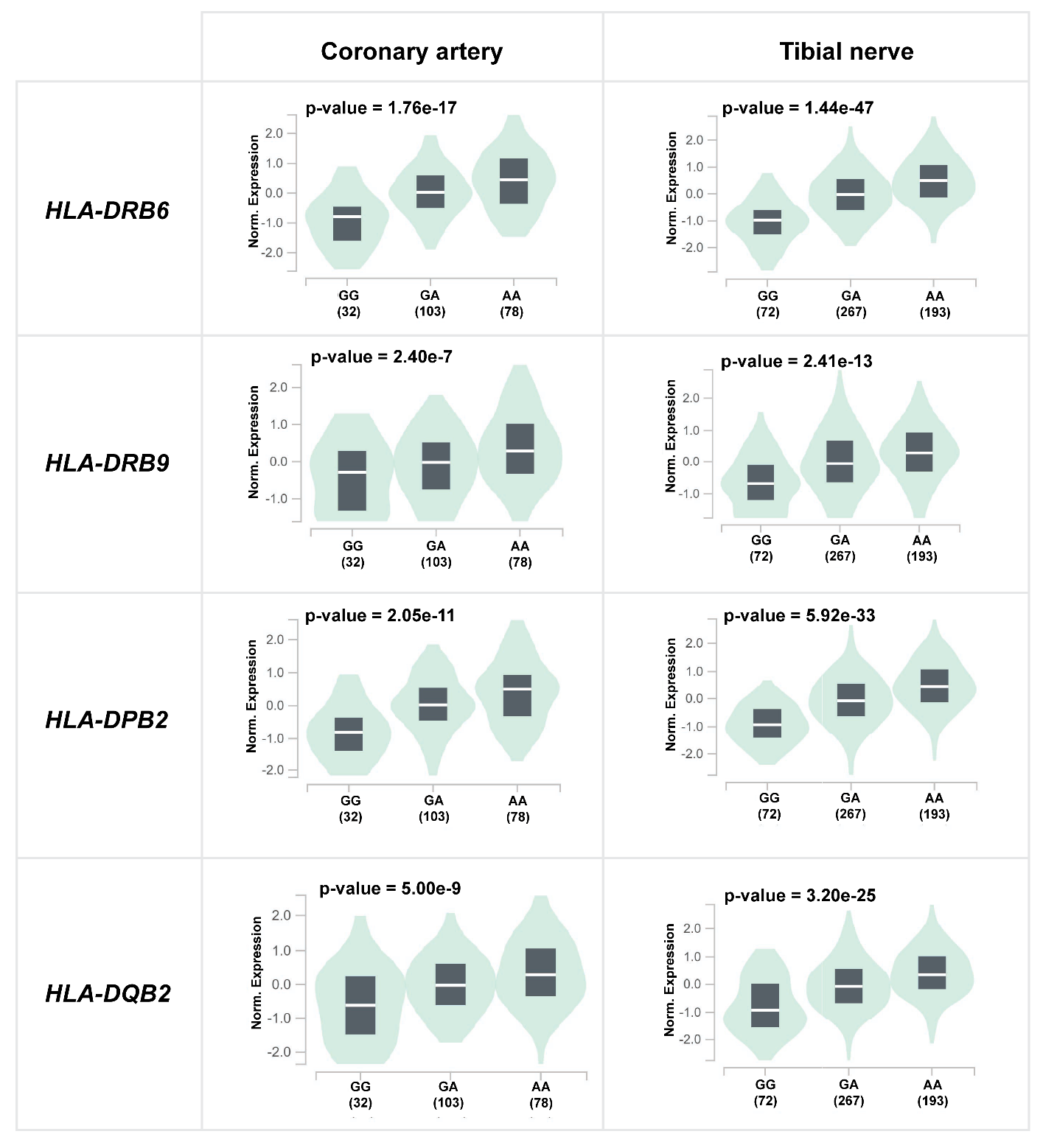

**Figure S6.** Genome browser view of H3K27ac ChIP-seq tracks in the surrounding region of rs9269041 (hg38, chr6: 32470466, red line) in CD14+ monocytes. Rows correspond to different ChIP-seq experiments from the ENCODE/Roadmap Epigenomics project databases. The upper panel displays a large region of chromosome 6 containing the *HLA-DRA* gene and the PAPS-associated SNP, while the lower panel represents a zoomed in region.

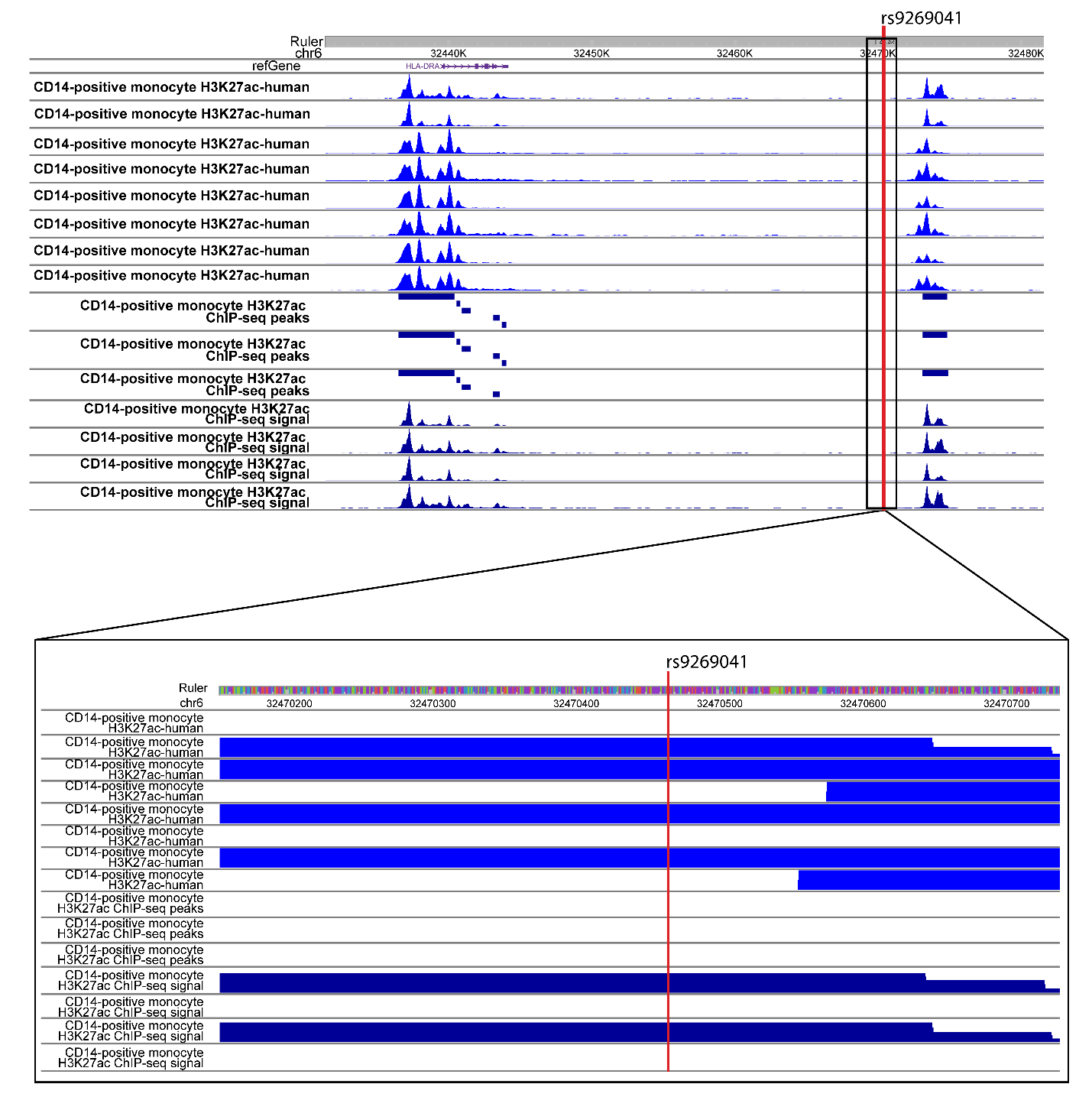
