## Supplementary material for "A genome-wide association study suggests new susceptibility loci for primary antiphospholipid syndrome": PRECISESADS Consortium

PRECISESADS Clinical Sites Contributors

Lorenzo Beretta^1^, Barbara Vigone^[1]^, Jacques-Olivier Pers^2^, Alain Saraux^2^, Valérie Devauchelle-Pensec^2^, Divi Cornec^2^, Sandrine Jousse-Joulin ^[2]^, Bernard Lauwerys^3^, Julie Ducreux^3^, Anne-Lise Maudoux^[3]^, Carlos Vasconcelos^4^, Ana Tavares^4^, Esmeralda Neves^4^, Raquel Faria ^[4]^, Mariana Brandão^4^, Ana Campar^4^, António Marinho^4^, Fátima Farinha^4^, Isabel Almeida^4^, Miguel Angel Gonzalez-Gay^5^, Ricardo Blanco Alonso^5^, Alfonso Corrales Martínez^[5]^, Ricard Cervera^6^, Ignasi Rodríguez-Pintó^6^, Gerard Espinosa ^[6]^, Rik Lories^7^, Ellen De Langhe^[7]^, Nicolas Hunzelmann^8^, Doreen Belz^[8]^, Torsten Witte^9^, Niklas Baerlecken ^[9]^, Georg Stummvoll^10^, Michael Zauner^10^, Michaela Lehner ^[10]^, Eduardo Collantes^11^, Rafaela Ortega-Castro^11^, Mª Angeles Aguirre-Zamorano^11^, Alejandro Escudero-Contreras^11^, Mª Carmen Castro-Villegas ^[11]^, Yolanda Jiménez Gómez^11^, Norberto Ortego^12^, María Concepción Fernández Roldán ^[12]^, Enrique Raya^13^, Inmaculada Jiménez Moleón ^[13]^, Enrique de Ramon^14^, Isabel Díaz Quintero^[14]^, Pier Luigi Meroni^15^, Maria Gerosa^15^, Tommaso Schioppo^15^, Carolina Artusi^[15]^, Carlo Chizzolini^16^, Aleksandra Dufour^16^, Donatienne Wynar^[16]^,

Laszló Kovács^17^, Attila Balog^17^, Magdolna Deák^17^, Márta Bocskai^17^, Sonja Dulic^17^, Gabriella Kádár^[17]^, Falk Hiepe^18^, Velia Gerl^18^, Silvia Thiel^[18]^, Manuel Rodriguez Maresca^19^, Antonio López-Berrio^19^, Rocío Aguilar-Quesada^19^, Héctor Navarro-Linares ^[19]^, Yiannis Ioannou^[20]^, Chris Chamberlain^20^, Jacqueline Marovac^20^, Marta Alarcón Riquelme^[21]^, Tania Gomes Anjos^21^.

1. Referral Center for Systemic Autoimmune Diseases, Fondazione IRCCS Ca’ Granda Ospedale Maggiore Policlinico di Milano, Italy.
2. Centre Hospitalier Universitaire de Brest, Hospital de la Cavale Blanche, Brest, France.
3. Pôle de pathologies rhumatismales systémiques et inflammatoires, Institut de Recherche Expérimentale et Clinique, Université catholique de Louvain, Brussels, Belgium.
4. Centro Hospitalar do Porto, Portugal.
5. Hospital Universitario Marqués de Valdecilla, IDIVAL, Universidad de Cantabria, Santander, Spain.
6. Hospital Clinic I Provicia, Institut d’Investigacions Biomèdiques August Pi i Sunyer, Barcelona, Spain.
7. Katholieke Universiteit Leuven, Belgium.
8. Klinikum der Universitaet zu Koeln, Cologne, Germany.
9. Medizinische Hochschule Hannover, Germany.
10. Medical University Vienna, Vienna, Austria.
11. Servicio Andaluz de Salud, Hospital Universitario Reina Sofía Córdoba, Spain.
12. Servicio Andaluz de Salud, Complejo hospitalario Universitario de Granada (Hospital Universitario San Cecilio), Spain.
13. Servicio Andaluz de Salud, Complejo hospitalario Universitario de Granada (Hospital Virgen de las Nieves), Spain.
14. Servicio Andaluz de Salud, Hospital Regional Universitario de Málaga, Spain
15. Università degli studi di Milano, Milan, Italy.
16. Hospitaux Universitaires de Genève, Switzerland.
17. University of Szeged, Szeged, Hungary.
18. Charite, Berlin, Germany.
19. Andalusian Public Health System Biobank, Granada, Spain
20. UCB Pharma, Slough, United Kingdom (PRECISESADS Project office)
21. Department of Medical Genomics, Center for Genomics and Oncological Research (GENYO), Granada, Spain (PRECISESADS Project Office)
